# Detecting SARS-CoV-2 Cryptic Lineages using Publicly Available Whole Genome Wastewater Sequencing Data

**DOI:** 10.1101/2024.12.24.24319568

**Authors:** Reinier Suarez, Devon A. Gregory, David A. Baker, Clayton Rushford, Torin Hunter, Nicholas R. Minor, Clayton Russ, Emma Copen, David H. O’Connor, Marc C. Johnson

## Abstract

Beginning in early 2021, unique and highly divergent lineages of SARS-CoV-2 were sporadically found in wastewater sewersheds using a sequencing strategy focused on the most mutagenic region of SARS-CoV-2, the receptor binding domain (RBD). Because these RBD sequences did not match known circulating strains and their source was not known, we termed them “cryptic lineages”. To date, more than 20 cryptic lineages have been identified using the RBD-focused sequencing strategy. Here, we identified and characterized additional cryptic lineages from SARS-CoV-2 wastewater sequences submitted to NCBI’s Sequence Read Archives (SRA). Wastewater sequence datasets were screened for individual sequence reads that contained combinations of mutations frequently found in cryptic lineages but not contemporary circulating lineages. Using this method, we identified 18 cryptic lineages that appeared in multiple samples from the same sewershed, including 12 that were not previously reported. Partial consensus sequences were generated for each cryptic lineage by extracting and mapping sequences containing cryptic-specific mutations. Surprisingly, seven of the mutations that appeared convergently in cryptic lineages were reversions to sequences that were highly conserved in SARS-CoV-2-related bat Sarbecoviruses. The apparent reversion to bat Sarbecovirus sequences suggests that SARS-CoV-2 adaptation to replicate efficiently in respiratory tissues preceded the COVID-19 pandemic.

**Author Summary:** Wastewater surveillance has been used during the SARS-CoV-2 pandemic to monitor viral activity and the spread of viral lineages. Occasionally, SARS-CoV-2 sequences from wastewater reveal unique evolutionary advanced lineages of SARS-CoV-2 from an unknown source, which are termed cryptic lineages. Many groups nationwide also use wastewater surveillance to track the virus and upload that information to NCBI’s SRA database. That sequence data was screened to identify 18 cryptic lineages worldwide and identify convergent mutations throughout the genome of multiple cryptic lineages that suggest reversion to residues common in SARS-CoV-2-related Sarbecoviruses.

## Introduction

Wastewater surveillance has been widely used to identify chemicals and microbes (1–3). During the SARS- CoV-2 pandemic, this technique gained prominence for its efficient tracking of various variants of concern (4). Our group began tracking SARS-CoV-2 lineages from wastewater in early 2021, and in March 2021, we discovered the first instance of an evolutionarily advanced SARS-CoV-2 receptor binding domain (RBD) haplotype that appeared repeatedly in a single sewershed, which we later termed a “cryptic lineage” (5). Examples of cryptic lineages have now been reported worldwide (5–11). Similarities between genomes from persistent SARS-CoV-2 infections in immunocompromised patients and cryptic lineages suggest these may reside within immunocompromised individuals (8,12,13). Furthermore, a single cryptic lineage derived from a lineage that stopped circulating in early 2021 was traced to a commercial building in late 2022, and 12S ribosomal RNA sequencing of the wastewater indicated that the only meaningful species contributing to the wastewater was human (13). Therefore, cryptic lineages are believed to be derived from individuals with very long SARS-CoV-2 infections.

Cryptic lineages often forecast mutations that are eventually acquired by circulating lineages. For instance, Spike substitutions N440K, S477N, E484A, and Y505H had not been seen in any major circulating lineages prior to Omicron. Yet, these mutations had repeatedly appeared in cryptic lineages long before Omicron emerged (5,6). The convergence between mutations found in cryptic lineages and those eventually found in circulating lineages suggests that cryptic lineages and major circulating lineages share selective pressures. However, many of the mutations seen repeatedly in cryptic lineages have yet to become prominent in any major circulating lineage (13). It is unknown whether major circulating lineages will eventually acquire those mutations or whether those mutations account for selective pressures that differ from circulating lineages.

Many organizations worldwide use whole genome sequencing (WGS) to detect and identify SARS-CoV-2 variants in wastewater samples. Much of this data is uploaded to the National Center for Biotechnology Information’s (NCBI) Sequencing Read Archive (SRA) or one of its international equivalents, the International Nucleotide Sequence Database Collection (INSDC). In this report, we screen 135,672 samples from over 2,000 sites across 45 countries and demonstrate the feasibility of screening the SRA database to detect SARS-CoV-2 cryptic lineages and analyze their mutations.

## Results

Using conservative thresholds, our lab has identified over 20 cryptic lineages by amplifying the RBD sequence from SARS-CoV-2 RNA in wastewater samples (5,6,13). From previously discovered cryptic lineages, we compiled a list of mutations observed in multiple cryptic lineages that had not yet been detected in any Omicron circulating lineage (S1 Figure). This list of 69 amino acid substitutions was termed “cryptic lineage-defining amino acid substitutions”

Using the search terms “SARS-CoV-2 wastewater”, we downloaded wastewater SARS-CoV-2 sequence reads from SRA that were available on February 18th, 2024, that had sample collection dates on or before October 31 2023, mapped these reads to the SARS-CoV-2 genome (NC_045512), and processed them with the program SAM Refiner (14). We identified individual sequencing reads in the SRA datasets that contained at least two of the cryptic lineage-defining amino acid substitutions (S1 Data). These were analyzed manually to identify haplotypes that did not match any known sequence from a patient sample and appeared multiple times in samples from the same sewershed. Using the subset of identified sequences, we found sequencing reads consistent with 18 independent cryptic lineages. Of the 18 identified lineages, three of the lineages we reported previously and three of the lineages had been reported by other groups (5–7,9,11,13). The duration of detection varied widely among the cryptic lineages; the shortest time a cryptic lineage was detected was one month (CA-1 and NY-2), while two cryptic lineages were detected for over a year (UK-1 and WI-1) (Table 1).

**Table 1.**
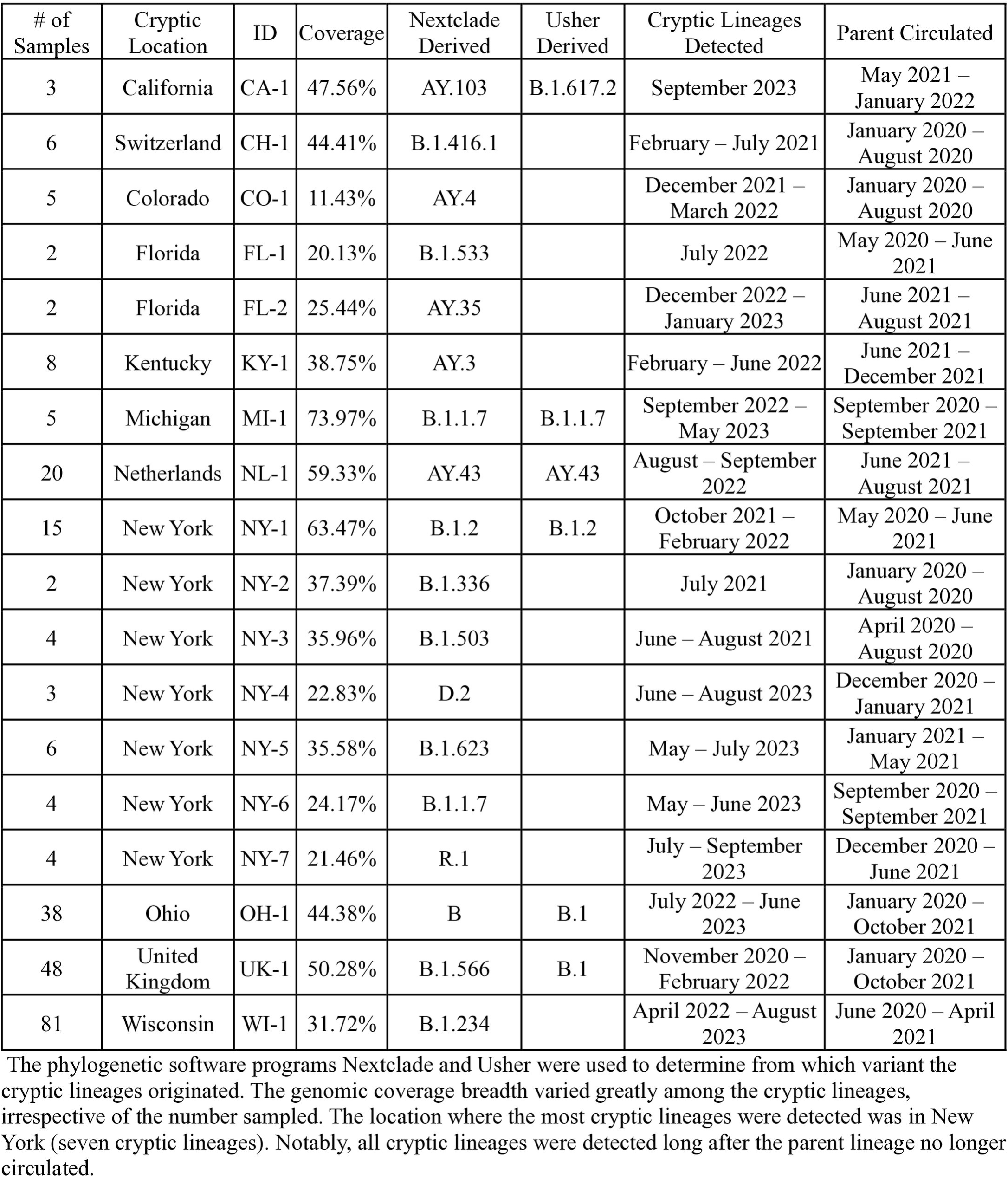
Total number of cryptic lineages found using our SRA screen.

After cryptic lineages are identified based on their RBD sequence, we can retrospectively identify other datasets from the same sewershed that share cryptic-defining characteristics outside of the RBD to partially reconstruct lineage genomes. We compared the individual SARS-CoV-2 sequences present in wastewater samples from sewersheds containing cryptic lineages to the sequences from samples from neighboring (same state) sewersheds collected during the same time period and, when possible, sequenced by the same agency (S2 and S3 Data). Individual mutations that appeared in multiple samples from the cryptic sewershed and were at least 50x more prevalent in the cryptic containing sewershed than in neighboring sewersheds were considered putatively cryptic-specific mutations (Figure 1a). Additionally, any mutation frequently appearing in the same sequence read as the cryptic-specific mutation was presumed to be present in the cryptic lineage (Figure 1b; see methods for specific criteria). This process was repeated with all 18 cryptic lineages to approximate the polymorphisms present in each lineage (S4 and S5 Data). A consensus sequence was generated for each cryptic lineage using its cryptic-specific mutations and sequences that appeared on the same read as the cryptic-specific mutations (Figure 1b, S6 Data File). Generating a complete consensus sequence for each cryptic lineage proved challenging. Sequence coverage varied between cryptic lineages, with the highest coverage being 73.97% (MI- 1) and the lowest 11.43% (CO-1). The consensus sequence was used as inputs for the phylogenetic software programs UShER (15) and Nextclade (16) to determine its predicted parent SARS-CoV-2 lineage (Table 1). All the cryptic lineages were predicted to be derived from lineages that stopped circulating months to years prior to their detection in wastewater (Table 1). A phylogenetic tree of the cryptic lineages illustrates the extreme diversity of these lineages (Figure 2). The use of a consensus sequence, which is derived from a mixture of diverse lineages with a shared common ancestor, could potentially influence the branch lengths in the phylogenetic tree, and may not fully capture the true diversity within each cryptic lineage.

**Fig 1.**
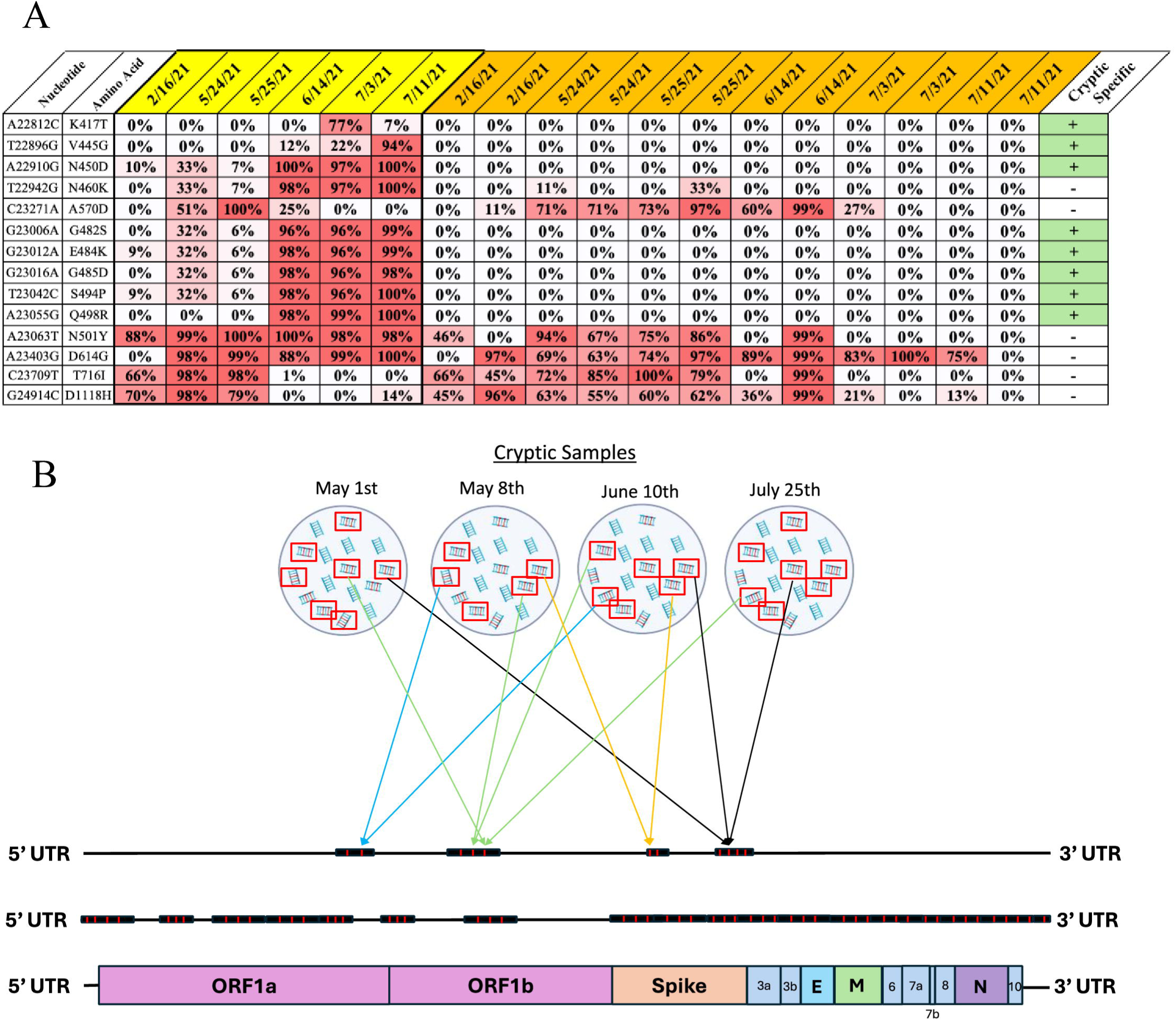
Schematic of workflow. Samples from sewer shed facilities containing cryptic lineages (yellow) were compared against samples from neighboring sewer sheds that did not contain cryptic lineages (orange). (A) Using the CH-1 cryptic lineage as an example, mutations found in at least two cryptic samples, with a prevalence of 50x more in the cryptic samples, are tentatively considered cryptic-specific (green). (B) The sequence reads containing cryptic-specific mutations (red box) were mapped onto the SARS-CoV-2 genome, with varying coverage across the genome to create a consensus sequence (middle genome). To be mapped onto the genome, a cryptic-specific sequence must appear in two or more samples.

**Fig 2.**
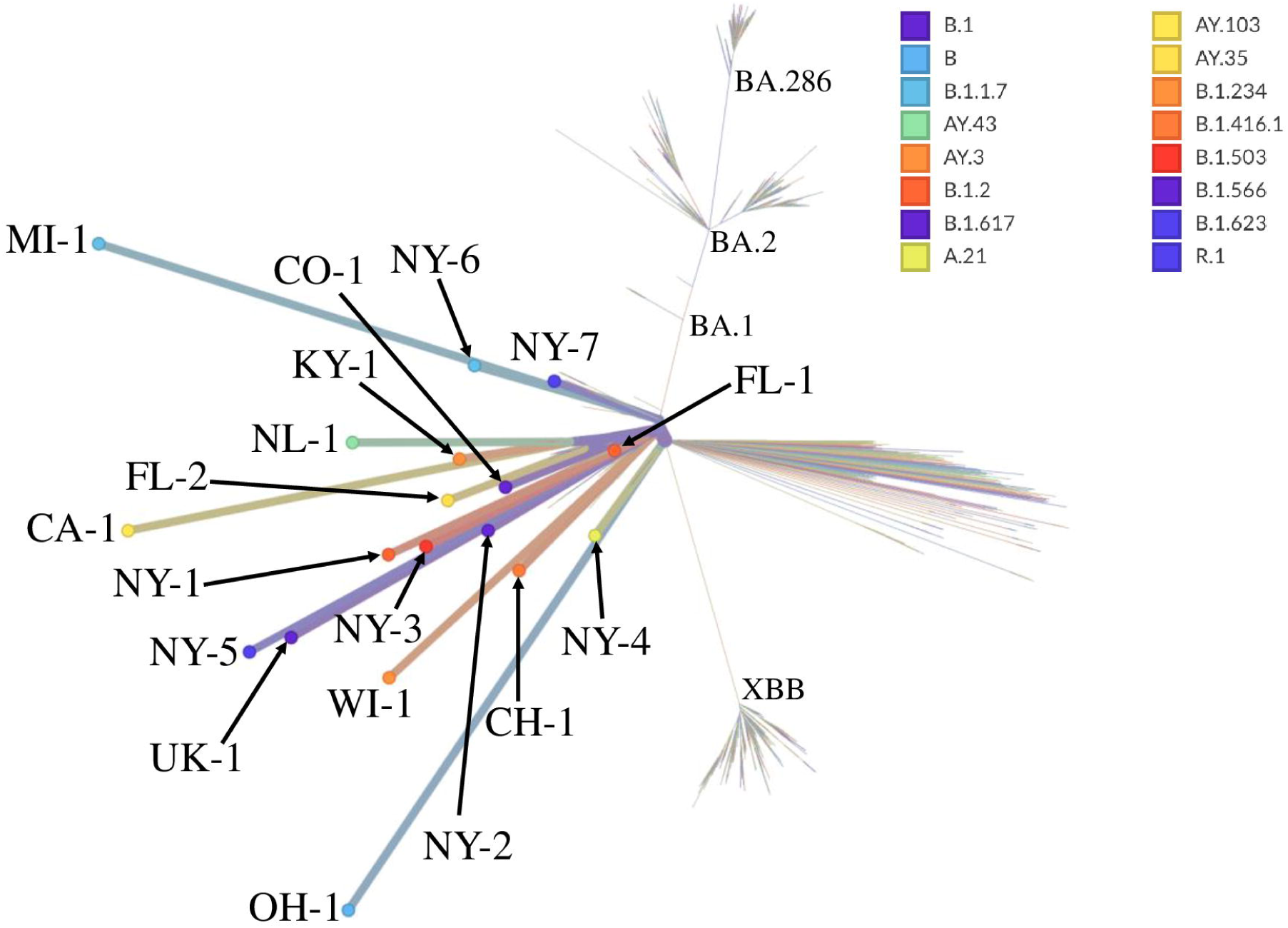
Generated phylogenetics tree using assemblies. The phylogenetic tree generated by NextClade illustrates the diversity of the cryptic lineages. The consensus sequences were uploaded onto Nextclade and compared against the Wuhan-Hu-1/2019 (MN908947). The hylogenetic tree highlights the diversity among the cryptic lineages detected.

Interestingly, we observed the same mutations in the consensus sequence appearing in multiple independent cryptic lineages. Such convergent changes are unlikely to be sequencing artifacts and likely reflect adaptation to common selective pressures. Mutations that appeared in three or more cryptic lineages were mapped onto a diagram of the SARS-CoV-2 genome while excluding mutations found in the parent lineages (Figures 3a & b; S7 Data). We observed 83 nucleotide changes in at least three cryptic lineages. The most common changes in Spike were K417T (78%) and Q493K (56%), which are known to affect antibody escape and ACE2 binding (17,18). Although K417T was present in the Gamma variant of concern and a few Omicron sub-lineages, such as BA.2.18, it has been present in less than 1% of circulating lineages found in people. By contrast, Q493K is extremely rare and has not been a lineage-defining change in any named PANGO lineage. The most common cryptic-specific mutations outside the Spike were in Orf1a (K1795Q) and Orf3 (H182D), each observed in 50% of the identified cryptic lineages.

**Fig 3.**
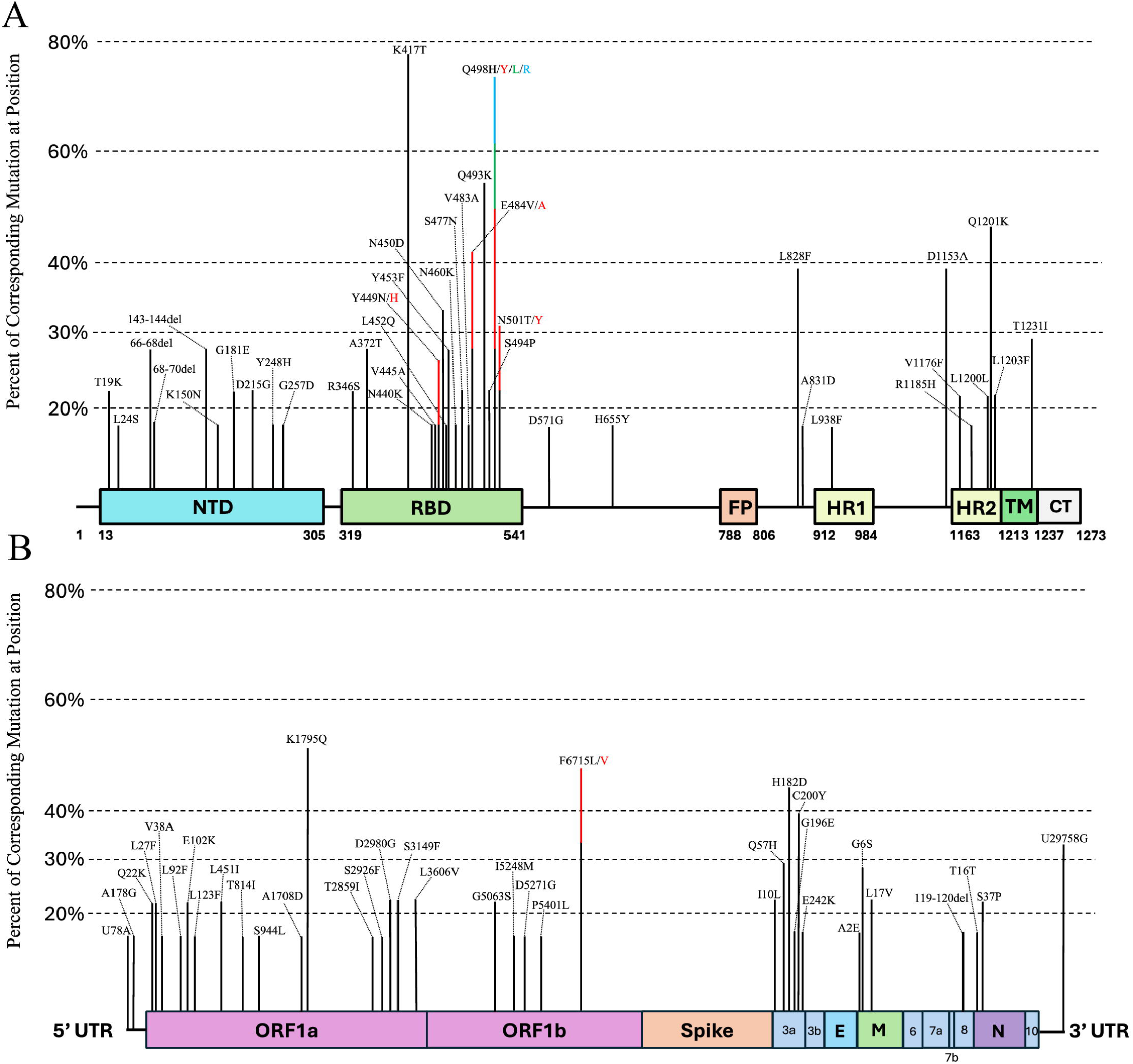
Assortment of stacked cryptic-specific mutations with a prevalence of ≥ 3. (A) Convergent mutations that appeared in at least three or more cryptic lineages were mapped onto the spike protein based on their location and prevalence across all the cryptic lineages. (B) Convergent mutations mapped against the SARS-CoV-2 genome, excluding the spike region. Positions which contain multiple mutations in the same position are represented as stacked bars and color coded.

Among the 83 changes that occurred convergently in at least three cryptic lineages, 79 changed a protein sequence through non-synonymous changes or deletions. Of the four changes that did not alter a protein sequence, two were silent (C25162A/Spike: L1200L, 22.22%), and three were in non-coding regions (T78A (16.67%), A178G (16.67%), and T29758G (33.33%)). Interestingly, we observed that the Spike change C25162A (L1200L) was always associated with the neighboring C25163A (Q1201K) change. These two mutations together create the sequence TCTAAAAGAACT, which is a near-perfect match to the consensus SARS-CoV-2 transcription regulatory sequence (TRS) TCTAAACGAACT (19). Although C25162A and C25163A are relatively rare in patient sequences, the two changes usually occur together (>60% of the time). While the function of this additional TRS is not known, it is a likely explanation for the convergence of the silent C25162A change.

A particularly notable convergent non-coding change in the cryptic lineages is at the 3’ UTR of the SARS-CoV- 2 genome, T29758G. This mutation is in the highly conserved region of the stem-loop two motif (s2m), which is found in many Coronaviruses and other RNA viruses (20–22). Remarkably, the s2m in SARS-CoV-2 deviates from the consensus s2m found in other RNA viruses, including Sarbecoviruses, and the T29758G mutation restores the SARS-CoV-2 to the consensus s2m sequence (22,23). The s2m stem-loop is not essential for replication as the sequence was deleted in omicron lineage BA.2 and all of its derivatives; thus, it has been nearly absent in circulating lineages for over two years (24). However, in the case of the cryptic lineages, the sequence frequently reverts the SARS-CoV-2 s2m to the Sarbecovirus consensus sequence.

Several of the most common convergent changes in cryptic lineages, such as Orf1a: K1795Q and T29758G, were conversions to the sequence found in closely related bat Sarbecoviruses such as RaTG-13. Although SARS-CoV-2 is a human respiratory pathogen, the most closely related Sarbecoviruses primarily infect Horseshoe bats and are primarily believed to be enteric pathogens. To explore if other convergent changes in cryptic lineages represent reversions to the Sarbecovirus consensus sequence, the sequences of seven closely related Sarbecoviruses (RpYN06, RATG-13, BANAL-52, BANAL-103, BANAL-116, and BANAL247) were compared to SARS-CoV-2 to identify amino acid positions that were conserved across all seven sarbecoviruses, but differed in the original SARS-CoV-2 A and B lineages. A total of 26 substitutions were identified where the SARS-CoV-2 sequence differed from all seven of the bat sarbecoviruses. Of these 26 positions, 12 substitutions in cryptic lineages had reverted to the Sarbecovirus consensus sequence in at least one cryptic lineage, and seven of the reversions occurred in at least three cryptic lineages (Figure 4). As of October 31st, 2023, in these 26 positions only one substitution that reverted to the Sarbecovirus sequence (ORF1a: A3143V) appeared in over 1% of all in-patient SARS-CoV-2 sequences. This high frequency of reversion to the consensus bat sarbecovirus sequence in cryptic lineages but not circulating lineages is consistent with cryptic lineages being subject to similar selective pressures as that of its bat progenitors.

**Fig 4.**
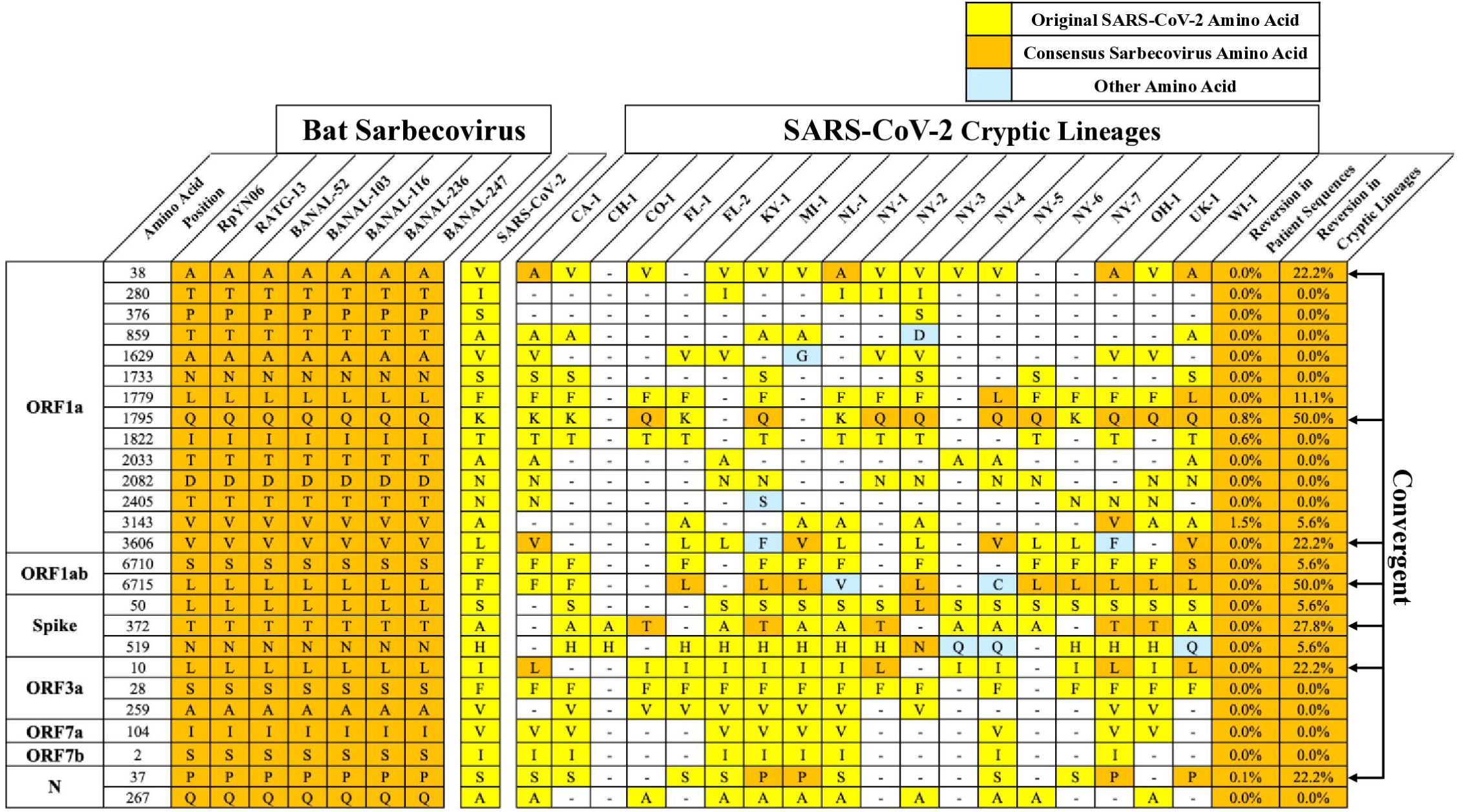
Chart of SARS-CoV-2 amino acids that deviate from the consensus Sarbecovirus amino acid sequence. Ubiquitous amino acids found across seven bat Sarbecoviruses (orange) highlight the occurrence of cryptic lineages to revert the SARS-CoV-2 (yellow) amino acid to the bat Sarbecovirus. The amino acid positions where a change is observed but differ from the Sarbecoviruses and SARS-CoV-2 are highlighted in blue. Instances where the same amino acid reversion occurred in ≥ 3 er tic lineages are designated as convergent.

Five of the cryptic lineages were found to have small insertions (Figure 5). Three of the insertions occurred in the ectodomain of the structural proteins, specifically in the spike and M genes, as was previously noted for one cryptic lineage, and the other two insertions occurred in non-structural genes, ORF3a and ORF7a (13). A closer observation of the inserted nucleotide sequence revealed that four of the five insertions were duplicated sequences from other parts of the SARS-CoV-2 genome.

**Fig 5.**
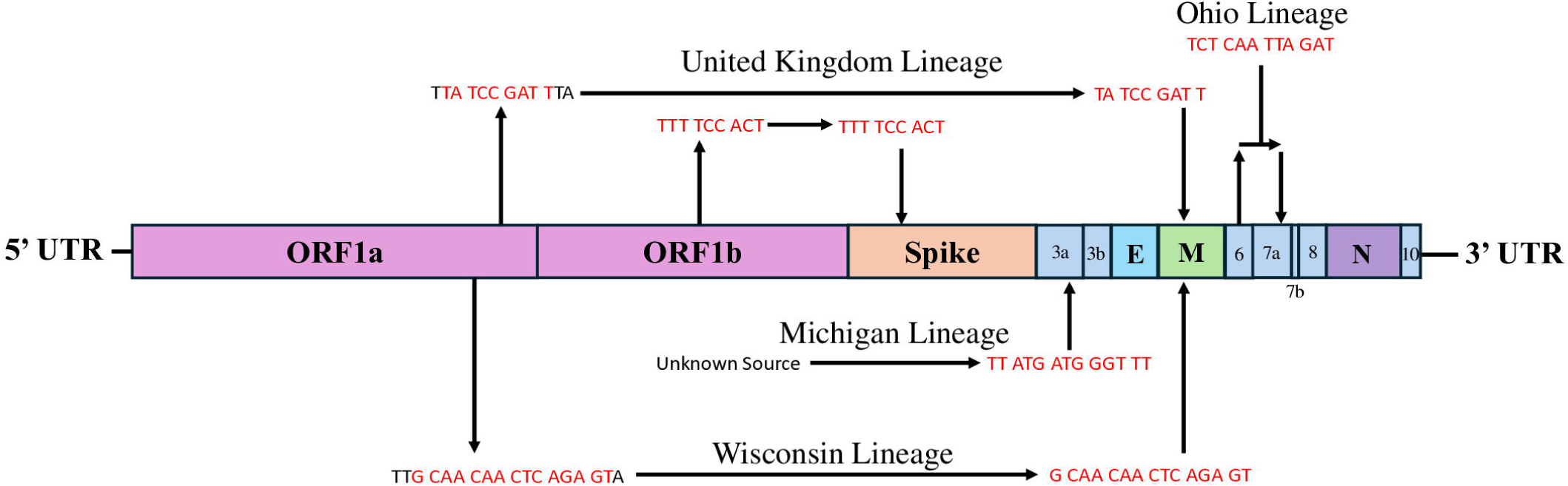
Insertion sequences were mainly derived from duplications. Insertion sites were mapped onto the SARS-CoV-2 genome to visually represent where the duplicated sequence (red) occurred and where the insertion was detected with respect to the cryptic lineage.

One cryptic lineage was detected in SRA datasets from two different sewersheds separated by approximately 40 miles. Samples were independently obtained from both sewersheds and tested for the presence of the cryptic lineage. Samples from both sewersheds contained a cryptic lineage that closely matched the sequence observed in the SRA sequences (S2 and S3 Figure). Similarly to the cryptic lineage found in Wisconsin (WI-1), the sequence from the Ohio cryptic lineage did not remain static over a nine-month period (Figure 6). Both sewersheds from Ohio shared highly similar cryptic-specific mutation profiles throughout the dates detected in the SRA. Notably, mutations in the Spike protein N460K, F486P, Q493T, and P499T were detected for the first time on the same date from both sewer sheds, strongly suggesting this lineage was being deposited into wastewater from a single source, likely a person that commuted between both locations. The Ohio cryptic lineage persisted until June 2023 before disappearing.

**Fig 6.**
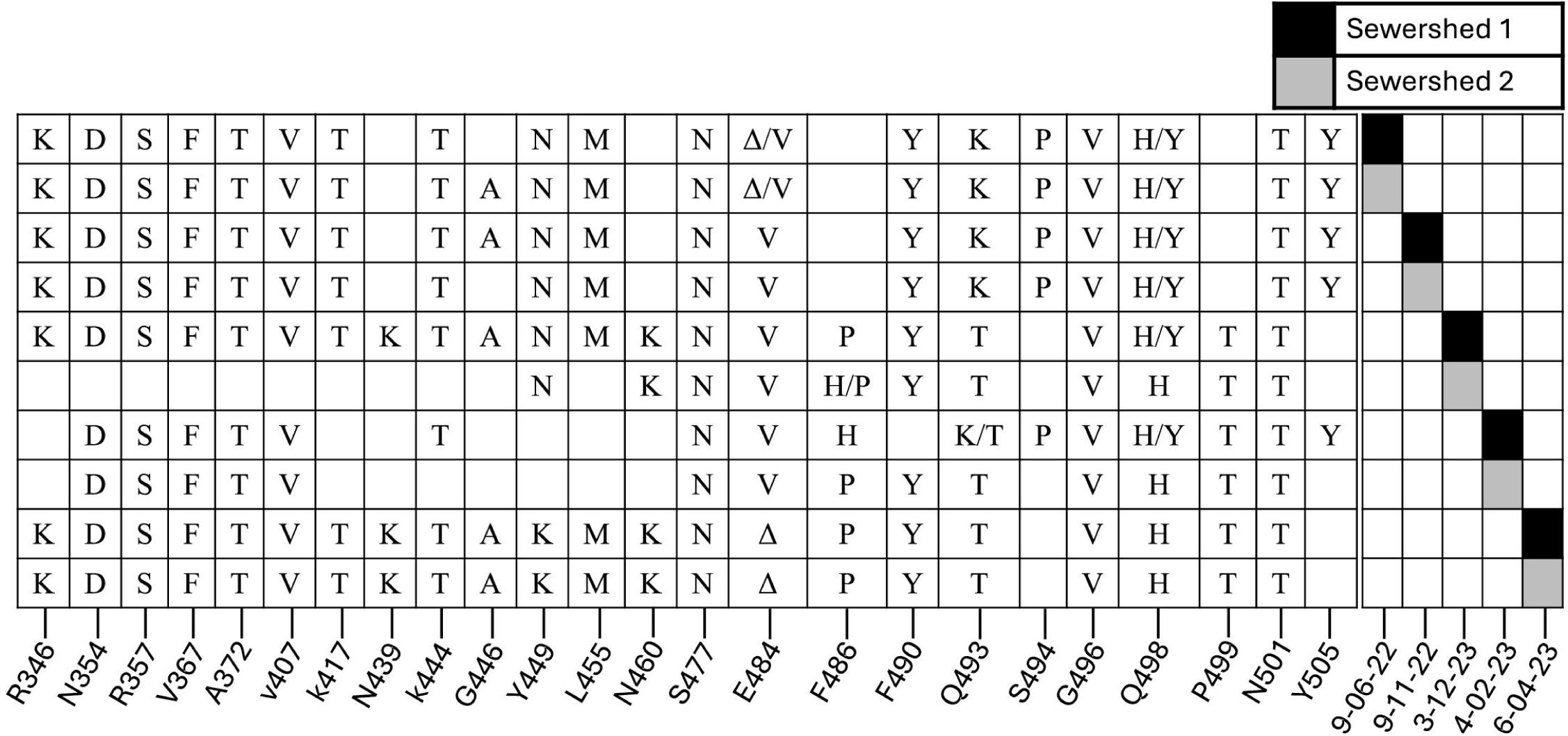
Cryptic-specific RBD mutations over time for the OH-1 cryptic lineage. Both locations shared highly similar mutation profiles in the RBD, with distinct mutations appearing in both locations around the same time (N460K, F486P, & P499T). Empty cells signify areas of low or no coverage.

## Discussion

Screening NCBI’s SRA database for cryptic lineages underestimates the prevalence of these lineages. Our screen relies on the detection of specific changes that are common to cryptic lineages, but there may be other cryptic lineages that do not harbor these conserved cryptic lineage signatures. Moreover, only a subset of global wastewater sequences are submitted to SRA, and the cryptic lineages need to be sufficiently abundant that their sequences can be detected after dilution with all of the other material in the sewershed. Despite these limitations, the method of cryptic lineage detection described here effectively detects cryptic lineages worldwide and highlights cryptic-specific polymorphisms outside the RBD. More importantly, this method illustrated cryptic-specific convergent polymorphisms across the many cryptic lineages.

Five insertion sites occur in various parts of the SARS-CoV-2 genome, but the impacts of these insertions are unknown. The insertions occurring in the structural regions of the SARS-CoV-2 genome (spike and M genes) are in the ectodomain section of the proteins. Studies have shown that SARS-CoV anti-M, in conjunction with anti-Spike, enhances the neutralizing capability of the virus (25–27). Thus, these insertions may contribute to immunological escape, while the significance of the escape requires testing. The role of the insertions in ORF3a and ORF7a are unknown; however, it is evident that SARS-CoV-2 readily utilizes the strategy of insertions as a form of adaptation to different selection pressures.

The K1795Q substitution is in the papain-like protease domain of nsp3 and the substitution has been shown to enhance the ability of the protease to cleave polyubiquitin chains (28). The most parsimonious explanation for the reversion of sequences in cryptic lineages to the sequence found in closely related Sarbecoviruses is that cryptic lineages are subject to selective pressures in common with enteric bat Sarbecoviruses that are not imposed on circulating lineages of SARS-CoV-2 that are primarily respiratory. The observation that enteric viruses consistently appear at >100 times higher levels than respiratory viruses in wastewater suggests that the digestive tract acts as a selective filter, diminishing much of the signal from respiratory viruses. This aligns with the observation that cryptic SARS-CoV-2 lineages, which are detected in wastewater and thought to originate from a single individual, are shed at extraordinarily high levels (13). The combined observations of cryptic lineages reverting to sequences found in their enteric ancestors, and their extremely high shed rates, are consistent with the idea that cryptic SARS-CoV-2 lineages predominantly replicate in the gastrointestinal (GI) tract.

The observation that SARS-CoV-2 contains at least seven distinct substitutions that convergently changed to the sequence found in enteric Sarbecoviruses suggests a strong conditional selective pressure to maintain the Sarbecovirus consensus sequence at these positions. The fact that SARS-CoV-2 had changes at each of these positions when it began circulating in humans suggests that SARS-CoV-2 had replicated in a non-enteric environment for a long enough period of time to allow these substitutions to persist and become fixed in the viral genomes that started the COVID-19 pandemic.

## Methods

### NCBI SRA Screening

All SARS-CoV-2 sequencing reads were obtained through the NCBI’s SRA and found by using the search terms “SARS-CoV-2 wastewater” then filtered to exclude any sample collected after October 2023. Raw reads were downloaded and mapped to the SARS-CoV-2 genome (NC_045512) using Minimap2 (29) and the resulting sam file processed by SAM Refiner with the parameters ‘—wgs 1—collect 0—indel 0—covar 0— min_count 1—min_samp_abund 0—min_col_abund 0—ntabund 0—ntcover 1’. Unique sequence outputs from SAM Refiner were programmatically screened for a combination of specific amino acid changes only found in cryptic lineages with positive hits manually validated. All scripts used in this study are publicly available through Github: https://github.com/dholab/SRA_wastewater_lineages.

### Cryptic-Specific Polymorphisms

To assess polymorphisms from sequence read runs (SRRs) containing cryptic lineages, we compared the sequences from sewer sheds containing cryptic lineages to sequences from neighboring (sewer sheds from the same state) sewer sheds that did not contain cryptic lineages. Two non-cryptic SRRs (negative samples) were compared against an SRR with a cryptic sequence. We selected negative and positive samples processed by the same sequencing agency to rule out testing bias. The selected SRRs were then processed using SAM Refiner, and the unique_seq and covar outputs were processed by a custom script to determine mutations associated with each cryptic lineage. The parameters for each cryptic-specific mutation are as follows: 1) The mutation must be present in SRA reads from two or more samples from a sewer shed where a cryptic lineage was observed; 2) the average sum abundance for the mutation must be 50x greater in the cryptic sewer sheds than in the non-cryptic sewer sheds; 3) a sum abundance of >10% of the maximum sum abundance of the most abundant polymorphism for a cryptic-specific mutation from those sewer shed samples. To account for mutations prevalent in both circulating and cryptic lineage, any polymorphism appearing at least 75% of the time in the same sequence read as a cryptic-specific polymorphism is considered part of the cryptic lineage and reported as “linked.”

The script generates three files for each cryptic lineage: a “CommonVars” file that lists all the polymorphisms found in all the samples compared (S3 Data File), a “Cryptic_CommonVars” file containing all the cryptic-specific mutations while flagging Delta, RaTG13, ubiquitous, and linked mutations (S4 Data File), and a “Cryptic_Covar” file that lists all the polymorphisms that were linked to cryptic-specific polymorphisms (S5 Data File). The cryptic-specific polymorphisms are then aggregated into a new file, “SortedVariance,” using a script that sorts them based on their prominence in all the cryptic lineages (S7 Data File). The cryptic-specific polymorphisms with a prevalence of ≥3 across all the cryptic lineages were then mapped onto a diagram of the SARS-CoV-2 genome based on their respective site.

### Ohio Cryptic Lineage Wastewater Sample Processing and RNA Extraction

24-hour composite samples of wastewater were collected weekly from the inflow of two undisclosed wastewater treatment facilities in Ohio. Samples arrived in 50mL conical tubes and were stored at 4°C until processed. Samples were centrifuged at 3000xg for 10 minutes and filtered through a 0.22μM polyethersulfone membrane (Millipore, Burlington, MA, USA). Approximately 37.5mL of wastewater was mixed with 12.5mL solution containing 50% (w/vol) polyethylene glycol 8000 and 1.2M NaCl, mixed, and incubated at 4°C. The samples would then be spun down at 12,000 RCF for 2 hours at 4°C. The supernatant was decanted, and the RNA was extracted from the remaining pellet using the QIAamp Viral RNA Mini Kit (Qiagen, Germantown, MD, USA) following the manufacturer’s instructions. The RNA was extracted in a final volume of 60uL.

### Amplifying the Ohio Cryptic Lineage

The primary RBD RT-PCR was conducted using the Superscript IV One-Step RT-PCR System (ThermoFisher Scientific, 12594100, Waltham, MA, USA). Primary RT-PCR amplification was performed as follows: [25 °C (2:00) + 50 °C (20:00) + 95 °C (2:00)] + ([95 °C (0:15) + 55 °C (0:30) + 72 °C (1:00)] × 25) cycles with the MiSeq primary PCR primers 5’-CAAACTTCTAACTTTAGAGTCCAACC-3’ and 5’- AAGTCCACAAACAGTTGCT-3’. An additional reaction was conducted to exclude omicron lineages utilizing the primer sets 5’-CCCTGATAAAGAACAGCAACC-3’ and 5’-TATATAATTCCGCATCATTTTCCAC-3’.

A secondary nested PCR (25μL) was performed on RBD amplifications using 5μL of the primary PCR as the template with MiSeq nested gene-specific primers containing 5′ adapter sequences (0.5μM each). The MiSeq nested RBD primer set for amplifying all lineage amplicons is 5’- gtgactggagttcagacgtgtgctcttccgatctACTACTACTCTGTATGGTTGGTAAC-3’ and 5’- acactctttccctacacgacgctcttccgatctCCTAATATTACAAACTTGTGCCCTT-3’. The MiSeq nested RBD primer set for amplifying excluded omicron amplicons is 5’- acactctttccctacacgacgctcttccgatctGTGATGAAGTCAGACAAATCGC-3’ and 5’- gtgactggagttcagacgtgtgctcttccgatctATGTCAAGAATCTCAAGTGTCTG-3’, along with dNTPs (100μM each) (New England Biolabs, N0447L) and Q5 DNA polymerase (New England Biolabs, M0541S, Ipswich, MA, USA). Secondary PCR amplification was executed as follows: 95 °C (2:00) + [95 °C (0:15) + 55 °C (0:30) + 72°C (1:00)] × 20 cycles. A tertiary PCR (50μL) was conducted to add the adapter sequences necessary for Illumina cluster generation using forward and reverse primers (0.2μM each), dNTPs (200μM each) (New England Biolabs, N0447L, Ipswich, MA, USA), and Phusion High-Fidelity or (KAPA HiFi for CA samples) DNA Polymerase (1U) (New England Biolabs, M0530L, Ipswich, MA, USA). PCR amplification was carried out as follows: 98 °C (3:00) + [98 °C (0:15) + 50 °C (0:30) + 72 °C (0:30)] × 7 cycles + 72 °C (7:00).

Amplified product (10μl) from each PCR reaction was combined and thoroughly mixed to create a single pool. The pooled amplicons were purified by adding Axygen AxyPrep MagPCR Clean-up beads (Corning, MAG- PCR-CL-50, Corning, NY, USA) at a 1:1 ratio to purify the final amplicons. The final amplicon library pool was evaluated using the Agilent Fragment Analyzer automated electrophoresis system (Agilent, Santa Clara, CA, USA), quantified using the Qubit HS dsDNA assay (ThermoFisher Scientific, Waltham, MA, USA), and diluted according to Illumina’s standard protocol. The Illumina MiSeq instrument generated paired-end 300 base pair reads (Illumina, San Diego, CA, USA). Adapter sequences were trimmed from the output sequences using Cutadapt.

Sequencing reads were processed as previously described (14). VSEARCH tools merged paired reads and dereplicated sequences (30). Dereplicated sequences from RBD amplicons were mapped to the reference sequence of SARS-CoV-2 (NC_045512.2) spike ORF using Minimap2 (29). Mapped amplicon sequences were then processed with SAM Refiner using the same spike sequence as a reference and the command line parameters “--Alpha 1.8 --foldab 0.6” (14). The haplotypes representing the Ohio lineages were rendered into figures using plotnine (https://plotnine.org).

### Phylogenetic Analysis

Phylogenetic trees were developed utilizing the software programs Nextclade (16) and UShER (15) using their default parameters. Each cryptic lineage had a consensus fasta file generated using the sequence reads containing cryptic-specific mutations (S5 Dataset). Non-cryptic specific mutations, which appeared at least 75% of the time in the same sequence read as a cryptic-specific mutation, are assumed to be part of the cryptic lineage and thus included in the consensus sequence. In positions where nucleotide mutations overlapped, the mutation with the highest abundance was chosen. If there was no coverage in a particular position or a mutation appeared ubiquitous in all samples, the designation “N” was used. To accurately generate the consensus sequence, only the last 35 positive cryptic lineage samples were used to create the consensus sequence. In Nextclade, consensus sequences were uploaded to the program, and each consensus was compared to the SARS-CoV-2 sequence (Wuhan-Hu-1/2019 (MN908947)). Using UShER, consensus sequences were copied onto the designated field and compared using the phylogenetic tree version “16,472,770 genomes from GISAID, GenBank, COG-UK and CNCB”.

## Supporting information

Description of Supplementary material

## Data Availability

All data produced in the present study are available upon reasonable request to the authors

