## Supplementary material for "Detecting SARS-CoV-2 Cryptic Lineages using Publicly Available Whole Genome Wastewater Sequencing Data": Description of Supplementary material

### **Description of Supplementary Data**

#### Supplementary Data 1 (S1_Data File): Screening of sequence reads that contained a combination of at least two polymorphisms from S1_Figure. This data lists the sequence read, its count and abundance, the SRR ID associated with the sequence, and the location from where it was sampled.

**Supplementary Data 2 (S2_Data File):** A list of all the samples (SRRs) screened in this study, along with their location and date sampled.

**Supplementary Data 3 (S3_Data File):** Referred to as “CommonVars”, this data file contains a list of all the polymorphisms found in both the cryptic positive SRRs (yellow) and cryptic negative SRRs (orange). This data highlights the count and abundance of each polymorphism in each sample and its sum in the positive, negative, and across all samples. To comply with submission guidelines, many low-abundance polymorphisms were excluded. A comprehensive report is available upon request from the corresponding author.

**Supplementary Data 4 (S4_Data File):** Referred to as “Cryptic_CommonVars”, this data file contains all the cryptic-specific polymorphisms found in the CommonVars data file. Additionally, this data highlights the count and abundance of each cryptic-specific polymorphism, in addition to flagging polymorphisms that appeared in the same sequence read as a cryptic-specific polymorphism (linked) and polymorphisms found abundantly in both positive and negative samples (ubiquitous). Additionally, this file flagged polymorphisms associated with the Delta SARS-CoV-2 lineage and the bat coronavirus, RATG13. Ubiquitous changes are noted but not included in the assemblies due to their ambiguity as belonging to the cryptic lineage. Yellow-highlighted SRRs indicate positive samples, while orange-highlighted SRRs indicate negative samples.

**Supplementary Data 5 (S5_Data File):** Referred to as “Cryptic_Covar”, this data file contains all linked polymorphisms. This data file highlights the count and abundance of linked polymorphisms and the SRRs in which they are found.

**Supplementary Data File 6 (S6_Data File):** List all fasta assemblies for all SARS-CoV-2 cryptic lineage identified. Regions or positions that displayed low or no coverage were considered ambiguous and designated with “N.”

**Supplementary Data File 7 (S7_Data File):** This data file is an assortment of all cryptic-specific polymorphisms across all the cryptic lineages.
