## Supplementary material for "Detecting SARS-CoV-2 Cryptic Lineages using Publicly Available Whole Genome Wastewater Sequencing Data": S1 Figure

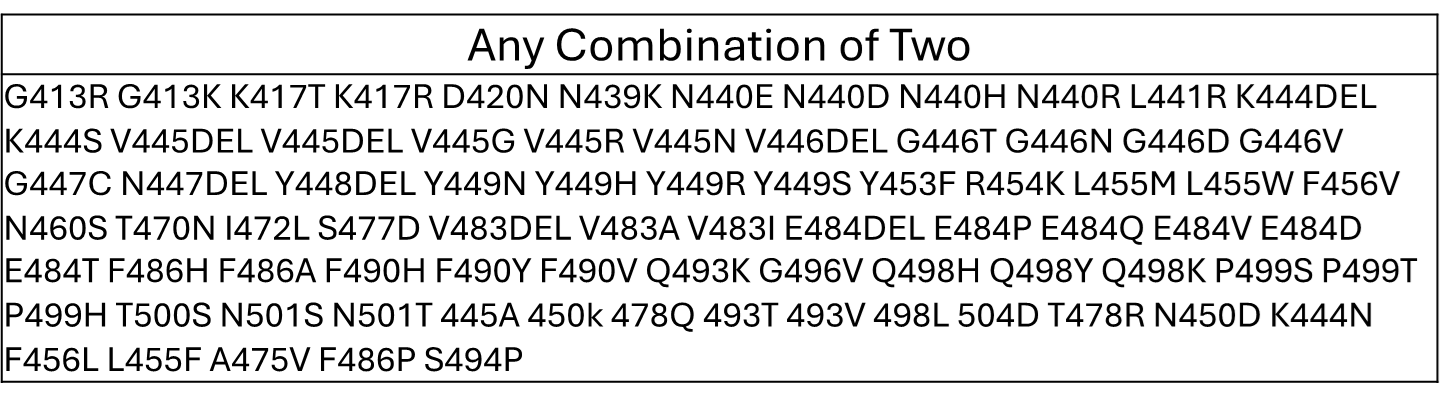
S Figure. Compiled list of mutations not found in major circulating lineages but frequently found in cryptic lineages.
