## Supplementary material for "Detecting SARS-CoV-2 Cryptic Lineages using Publicly Available Whole Genome Wastewater Sequencing Data": S2 Figure

S2 Figure. SARS-CoV-2 haplotype from the second location in Ohio.

RBD-focused amplifications of samples collected from the second location in Ohio. Amplifications using the omicron exclusion primer sets are designated as ALT.
