## Supplementary material for "Detecting SARS-CoV-2 Cryptic Lineages using Publicly Available Whole Genome Wastewater Sequencing Data": S3 Figure

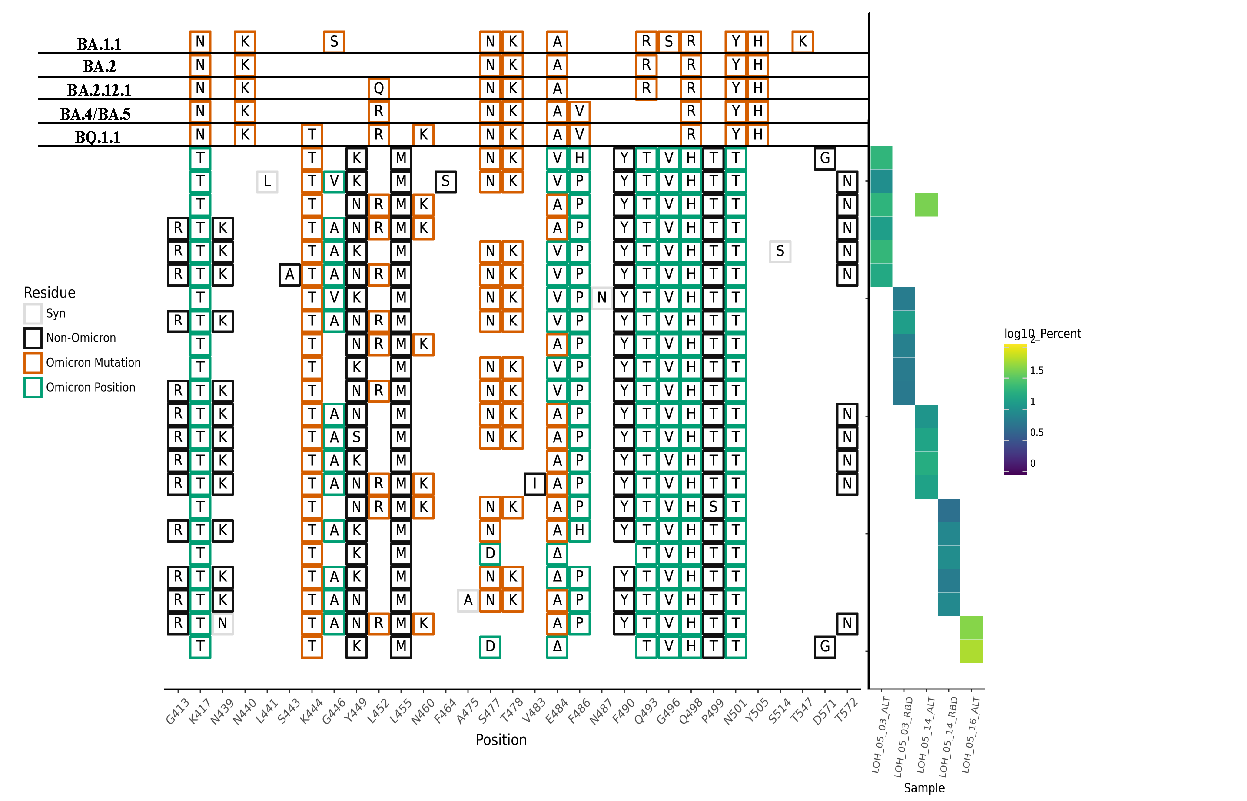

S3 Figure. SARS-CoV-2 haplotype from the first Ohio location.

RBD-focused amplifications of samples collected from the first location in Ohio. Amplifications using the omicron exclusion primer sets are designated as ALT.
